# Phenotypic models of drug-drug-gene interactions mediated by cytochrome drug-metabolizing enzymes

**DOI:** 10.1101/2023.11.02.23297749

**Authors:** Roberto Viviani, Judith Berres, Julia C. Stingl

## Abstract

Genetic polymorphisms in drug metabolizing enzymes and drug-drug interactions are major sources of inadequate drug exposure and ensuing adverse effects or insufficient responses. The current challenge in assessing drug-drug gene interactions (DDGI) for the development of precise dose adjustment recommendation systems is to take into account both simultaneously. Here, we analyze the static models of DDGI from *in vivo* data and focus on the concept of phenoconversion to model inhibition and genetic polymorphisms jointly. These models are applicable to datasets where pharmacokinetic information is missing and are being used in clinical support systems and consensus dose adjustment guidelines. We show that all such models can be handled by the same formal framework, and that models that differ at first sight are all versions of the same linear phenoconversion model. This model includes the linear pharmacogenetic and inhibition models as special cases. We highlight present challenges in this endeavour and the open issues for future research in developing DDGI models for recommendation systems.

## Introduction

Drug interactions and the ensuing side effects are an important clinical and public health issue. Major reasons of side effects are individual variability in enzymatic drug clearance due to genetic polymorphisms in drug metabolizing enzymes (DMEs, Stingl et al. 2013), and to drug-drug interactions (Tornio et al. 2019) causing changes in drug metabolism. In patients treated for depression, such changes may affect up to 25% of patients or more, highlighting the importance of this issue in personalizing treatment (Preskorn et al. 2013; Gloor et al. 2022; see also Mostafa et al. 2021).

Genetic polymorphisms in cytochrome DMEs such as CYP2C9, CYP2C19 and CYP2D6 lead to altered enzymatic activity that affects the metabolism of about one third of drugs on the market, especially psychoactive drugs (Ingelman-Sundberg 2004). Quantitative estimates of clearance changes for individual drug dosing (Kirchheiner et al. 2001, 2004; Stingl et al. 2013) and consensus-guided recommendations (such as caution in carriers of a genetic variant and a susceptible medication) have long been available (for an overview of current guidelines, see Abdullar-Koolmes et al. 2021). A recent study has shown that the systematic adoption of a pharmacogenetic recommendation system in the clinic may lead to a 30% reduction in adverse drug reactions relative to usual clinical practice (Swen et al. 2023). We will refer here to carriers of (wildtype) alleles associated with typical enzyme activity as NMs (normal metabolizers), and carriers of variants with no activity as PMs (poor metabolizers).

Drug-drug interactions may arise through diverse mechanisms, but an important one arises from increased drug exposure following inhibition of DME activity (*inhibitors*). Since characterization of inhibiting capacity of DME is mandatory to obtain market authorization approval from regulators, specific warnings on drug interactions are included in drug labels and are included in clinical decision support systems.

The challenge for the next generation of recommendation systems consists in using information on genetic variants and on drug-drug interactions (such as those ensuing from inhibition) simultaneously. The interplay between these two sources of changes in drug exposure has long been recognized and termed drug-drug gene interactions (DDGI; for reviews, see Bahar et al. 2017; Storelli et al. 2018). There is a wide consensus in the field that the extent of the inhibition may vary depending on the phenotype (Cicali et al. 2021), but different methods are in use to combine pharmacogenetic effects and inhibition. Our purpose here is to review the approaches to jointly model pharmacogenetic effects and inhibition, a key issue to predict DDGI.

We focus here on static models estimated entirely from *in vivo* data, including those obtained in clinical settings such as therapeutic drug monitoring (TDM). Such data do not provide information that enables estimating pharmacokinetic curves, as in pharmacokinetic models of DDGI (Vieira et al. 2014a; Storelli et al. 2019). Static models have the advantage of allowing us the systematic use of a wide range of *in vivo* data, including clinical data, in a meta-analytic framework. Furthermore, by covering many substances and DMEs at once, these models are being incorporated in recommendation systems and in consensus guidelines. We refer to this static modelling approach as *phenotypic modelling*.

One possible starting point for static models of DDGIs is the notion, formalized also in regulatory guidance (U.S. Department of Health and Human Services Food and Drug Administration 2020), that in real-world assessments changes in substrate exposure may be all viewed from the standpoint of changes in DME activity (Gibbs et al. 2006). Enzyme inhibition from an interacting drug may be characterized as *phenoconversion* to a phenotype with lower enzymatic activity. For example, complete inhibition of a DME in an NM individual may be described as phenoconversion from NM to the PM phenotype. Hence, an important task for phenotypic models of joint effects of pharmacogenetic variants and drug-drug interactions is to verify empirically that joint modelling can be based on this notion.

Before introducing models of phenoconversion, we will preliminarily examine existing phenotypic models of effects of genetic polymorphisms on drug metabolism and of inhibition from drug-drug interactions, taken separately. These are potentially very wide fields, and our exposition will necessarily be limited to the aspects that are relevant to formulate the models of joint effects from *in vivo* data that follow. Importantly, we will show that a common formal framework unifies all phenotypic models of pharmacogenetic effects and inhibition. This may justify its extension to joint models of both pharmacogenetic and inhibition effects on drug metabolism. Our purpose will be to express the variety of existing approaches in modelling phenoconversion and DDGIs through this unified framework. A list of used abbreviations is at the end of the main text.

## Results

### Phenotypic models of pharmacogenetic effects on metabolism

Early approaches to estimate effects of pharmacogenetic effects on drug exposure were based on computing separate averages of drug clearance measurements in every combination of phenotype and drugs (Kirchheiner et al. 2001, 2004; Stingl et al. 2013). While intuitive and easily interpretable, these models made no use of knowledge about the relationship between measurements of the same drug and phenotype, with possible negative consequences on the efficiency of estimates and their generalization capacity.

An insight emerging from this work was that when the effects of genetic polymorphisms are assessed on clearance ratios of the variant phenotype and the wild-type normal metabolizer type the resulting relation is at least approximately linear. This means that, if the activity scores of an allelic combination are known, one can use linear regression to estimate the predicted clearance ratio from the activity score *AS* (Stingl et al. 2022). Activity scores, introduced by Gaedigk et al. (2008) as estimates of DME activity predicted from the genotype, assign a value of zero to the PM phenotype (no metabolic capacity) and increasing values to phenotypes with increasing capacity. Activity scores model only the genetic variants that translate into changes in enzyme capacity.

To aid intuition in showing the relationship between pharmacogenetic effects and linear regression, it may be useful to introduce *relative activity scores RAS*, which are activity scores rebased such that the value for NMs is zero and positive and negative scores reflect increased or decreased enzyme activity: *RAS = AS* − *AS*_NM_, *AS = RAS* − *RAS*_PM_ (here, *RAS*_PM_ is the relative activity score of PMs and *AS*_NM_ the activity score of NMs). With this substitution, the relationship between clearance and enzyme activity may be expressed in its simplest form,

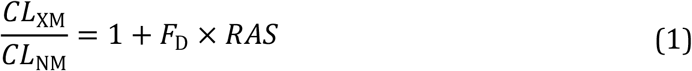

Here, 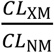 is the clearance ratio between carriers of a variant phenotype XM and NM individuals with typical enzyme activity, as reported in pharmacogenetic studies. In the right term, *F*_D_ represents the importance of the DME in question in the metabolism of a specific drug *D*, which also determines the extent to which polymorphism of the DME affect its clearance. Negative *RAS* values, representing reduced metabolic capacity, lead to negative change estimates in drug clearance relative to the baseline value of unity, which refers to the clearance ratio of normal metabolizers NMs. Changes in the opposite directions ensue from positive *RAS* values. Given *RAS*, a linear regression with a fixed intercept may be used to estimate the coefficient *F*_D_, which gives the slope of the straight line representing the relationship between activity scores and predicted clearance ratios (see Figure 1A for an example).

**Figure 1.**
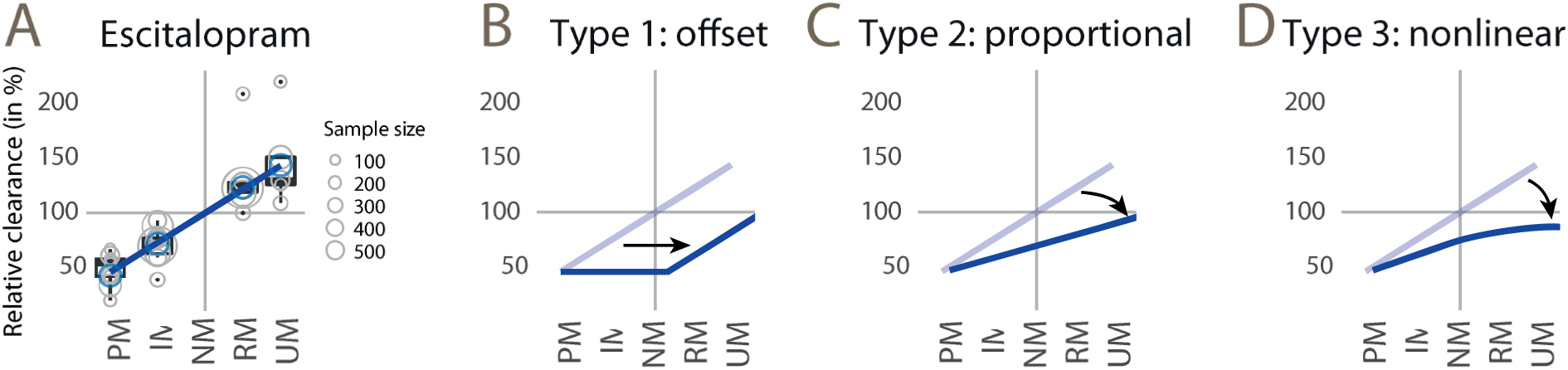
A: example of a regression of clearance ratio data from pharmacogenetic studies of escitalopram on relative activity scores (expressed on the *x* axis by the CYP2C19 phenotype). The slope of the fitted straight line represents the importance of the pathway of a polymorphic DME in the metabolism of this drug. The radius of the circles is the sample size of individual datapoints (from Stingl et al. 2022). B: the offset phenoconversion model is equivalent to a horizontal shift of this straight line to the right to an extent representing inhibition, bounded below by the adjustment for the PM phenotype. C: the proportional model uses a multiplicative coefficient to change the slope of this line. D: the non-linear model gives a more flexible prediction of phenoconversion. PM, IM, NM, RM, UM: poor, intermediate, normal, rapid, and ultrarapid metabolizer phenotypes.

To better understand *F*_D_, let us scale the predictor variable in this regression, i.e. the relative activity scores, such that *RAS*_PM_ *=* −1. Solving for *F*_D_ in PMs, one obtains

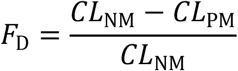

From this, we see that with this scaling of activity scores, *F*_D_ becomes an estimate of the fractional contribution of the DME to the clearance of drug *D* from *in vivo* data.

Relative to the earlier models, this approach offers the advantage that only one coefficient per drug is estimated by linear regression from the information from all phenotype groups, whose data are pooled in the estimate to gain efficiency. After the fit, predictions of changes in clearance ratios may be computed also for allelic combinations for which data are missing if the activity scores are known. Modelling all drugs together also allows testing effects of the types of studies in the database and verify that they do not lead to systematic bias.

While this model uncovers the relationship between activity scores and DME activity in its simplest form, it was not the first to be proposed in the literature. Tod et al. (2011) may have been the first to propose a static model of pharmacogenetic effects of CYP2D6 polymorphism on drug metabolism using a formalism that appears to differ from the one presented here. However, expressed in terms of clearance ratios, the model by Tod et al. may be shown to be equivalent to the model of eq. 1 (for details of the model and proof, see Appendix A1). Both models assume that the fractional contribution of the DME in the metabolic pathway of the drug remains the same for different phenotypes. This assumption is not likely to hold if a DME is the primary metabolizer of a drugs in NMs, as other DMEs may take over metabolism in PMs. However, for the majority of drugs linearity can be verified empirically (Stingl et al. 2022). Furthermore, both models assume that all polymorphic alleles and their activity scores are known.

When appropriate, dose adjustments may be based directly on estimated changes in metabolic clearance (i.e., a 50% fractional reduction in clearance may be compensated by a 50% reduction in doses; Stingl et al. 2013). This remains the case for all models that follow and estimate changes in clearance ratios.

### Static models of inhibition from *in vivo* data

Models of inhibition from *in vivo* data adapted mechanistic static models, i.e. static models predicting changes in metabolism from *in vitro* data (for comprehensive treatments, see Fahmi et al. 2009; Guest et al. 2011; Vieira et al. 2014b). One such model was proposed by Ohno et al. (2007) for the inhibition of CYP3A4, based on several simplifying assumptions, including equal intestinal availabilities:

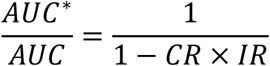

where the asterisk denotes the AUC in the inhibited condition, *CR* is the fraction contributed by the DME in the hepatic metabolism of the drug, and *IR* is an inhibition rate ranging from zero (no inhibition) to unity (full inhibition) and is defined by *IR = I*_app_⁄(*I*_app_ + *K*_*i*_), where *I*_app_ is the apparent time-averaged liver concentration of the inhibitor, and *K*_*i*_ a inhibitor-specific inhibition constant determined *in vitro*. Instead of estimating *IR* from *in vitro* data predicting the hepatic concentrations from dosage, Ohno et al. (2007) observed that *CR* and *IR* can estimated from *in vivo* AUC or clearance data when either one of these values is known. These values are readily available when the dose of the inhibitor is large enough to be regarded as exerting the maximal inhibition that can be obtained with that inhibitor. Therefore, they bootstrapped their analysis by setting *CR* to unity in a selective CYP3A4 substrate and progressively deriving *CR* and *IR* from a database of studies of drug-drug interactions. Departing from other mechanistic static models, Ohno et al. (2007) argued that *IR*, estimated from *in vivo* data, may be an acceptable index of inhibition irrespective of the mechanism through which inhibition ensues. Similarly, other sources of variation in the prediction from *in vitro* to *in vivo* inhibition may be overcame by using *in vivo* data directly. Over the years, this model was updated with the progressive availability of more data on drug interactions (Hisaka et al. 2010a, b; Loue and Tod 2014; Gabriel et al. 2016; Di Paolo et al. 2021, 2022). Tod et al. (2016) showed that the fit of this model compared favourably with that of the PBPK approach.

As noted by Ohno et al. (2007), in selective inhibitors of polymorphic DMEs *CR* may also be estimated from pharmacogenetic studies, since in full inhibition as well as in PMs the activity of the DME is zero. This relation may be used to derive an equivalent expression of the model by Ohno et al. (2007) in terms of the pharmacogenetic model of clearance ratios (eq. 1) introduced in the previous section (for derivation, see Appendix 2):

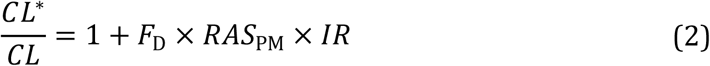

This model of inhibition takes the same linear form as the pharmacogenetic model (eq. 1, with *RAS*_PM_ × *IR* replacing *RAS*). Intuitively, it expresses the inhibition as a fraction of the maximal reduction in clearance as one may observe in the PM phenotype. With some terminological abuse, this may be viewed as a phenotypic model of inhibition in the sense that it characterizes a single quantity observable *in vivo* irrespective of the mechanisms that determine it.

The original aim of drug interaction models was to predict *in vivo* from *in vitro* interaction data, which may be relevant for example for drug development. For the purposes of drug development, it is more important to establish the very existence of a potential interaction than quantify it depending on the dose of the inhibitor. For this reason, although *IR* can in principle express the effects of the dose of the inhibitor (Tod et al. 2013), these models have used mostly data from studies of high doses of the inhibitor, which are obviously more appropriate to establish if an inhibitor has an effect on a victim drug at all. Static inhibition models from *in vivo* data may need further validation studies to understand their applicability to predict extent of inhibition at different doses of the inhibitor, as they may be used in the clinic.

### Phenotypic models of phenoconversion

A seminal work by Borges and colleagues (Borges et al. 2006) provided evidence that drug-drug inhibition differs depending on the DME activity predicted by *CYP2D6* genetic variants. This work and its sequel (Borges et al. 2010) were at the origin of the most common approach to combining information on pharmacogenetic variants and inhibitors (see the discussion by Cicali et al. 2021). Before discussing phenoconversion models that followed this work, we will introduce here a formal specification of possible alternatives.

In the phenotypic modelling framework described above, phenoconversion may be modelled by a function *f*(*IR, RAS*) which, given the *RAS* as predicted by the allelic composition of the individual (here referred to as the *genetypic RAS*) and IR, representing the inhibition arising from an inhibitor, returns a *phenoconverted activity score* that may be used in the place of *RAS* in the pharmacogenetic model of eq. 1:

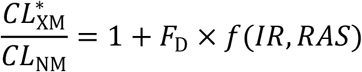

Note that the phenoconversion function makes no use of information on how *F*_D_ is determined. This is an important assumption, as it means that data from the importance of a DME in the metabolic pathway of a drug (represented by *F*_D_) and data on the capacity of the inhibitor contribute independently to the changes in clearance. All phenoconversion models in the literature make this assumption. As a further consequence, the model remains linear in the phenoconverted activity score. Hence, under this assumption the remaining issue concerns the form of the phenoconversion function *f*. There are several conceivable possibilities.

1. The first is to obtain the phenoconverted activity score by subtracting a quantity related to inhibition to the genotypic *RAS*, bounded below at the PM phenotype. This is equivalent to shifting the line predicting the dose recommendation by a fixed offset to the right (Figure 1B, *offset model*). This bound introduces a non-linearity.
2. A second possibility is that *IR* affects the genotypic *RAS* by a multiplicative factor. In this model, the slope of the line estimating the dose adjustment is modified by inhibition (Figure 1C, *proportional model*). By adjusting *RAS* multiplicatively, this model allows for the effect of inhibition to differ in the UM allelic configuration and in lower activity configurations while maintaining linearity.
3. A third possibility is that *IR* and the genotypic *RAS* interact in determining phenoconversion, or that there is an additional offset term, again bounded below by the PM phenotype. This would lead to a non-linear relationship between the genotypic *RAS* and the predicted clearance ratio (Figure 1D, *nonlinear model*).

### Models and evidence on phenoconversion in the literature

A summary of phenoconversion models is given in Table 1, together with existing clinical support systems. Perhaps because of their apparent simplicity, offset models are well represented in the literature. The first report of DDGI described, without formalizing it, data compatible with an offset model (Borges et al. 2006). An offset model was used by Mostafa et al. (2019) to evaluate the potential impact of DDGIs in actionable variants in an Australian sample. Bousman et al. (2021) shift the phenotype down to the next lower phenotype activity for moderate inhibitors, and to PM in strong inhibitors.

**Table 1.**
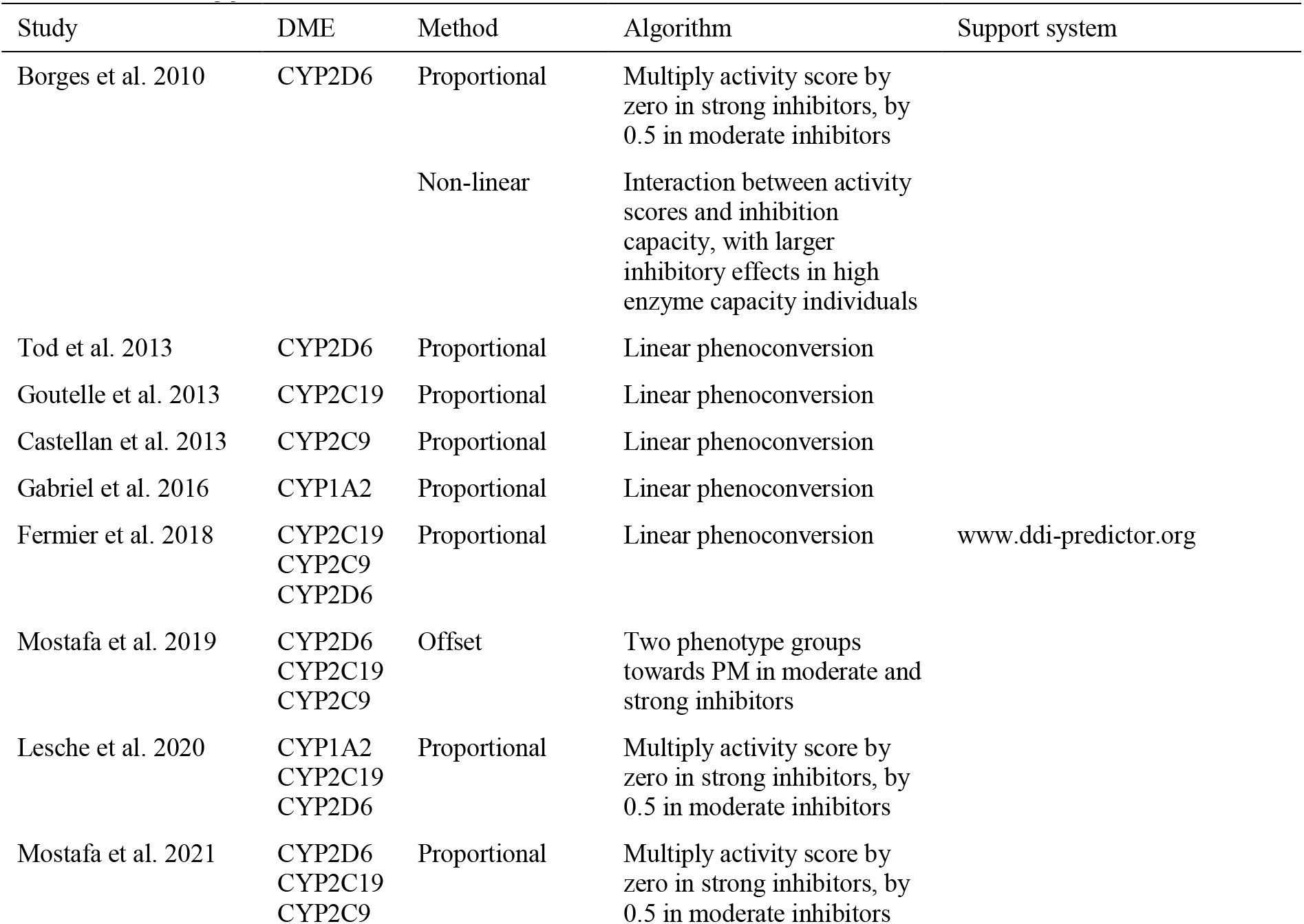
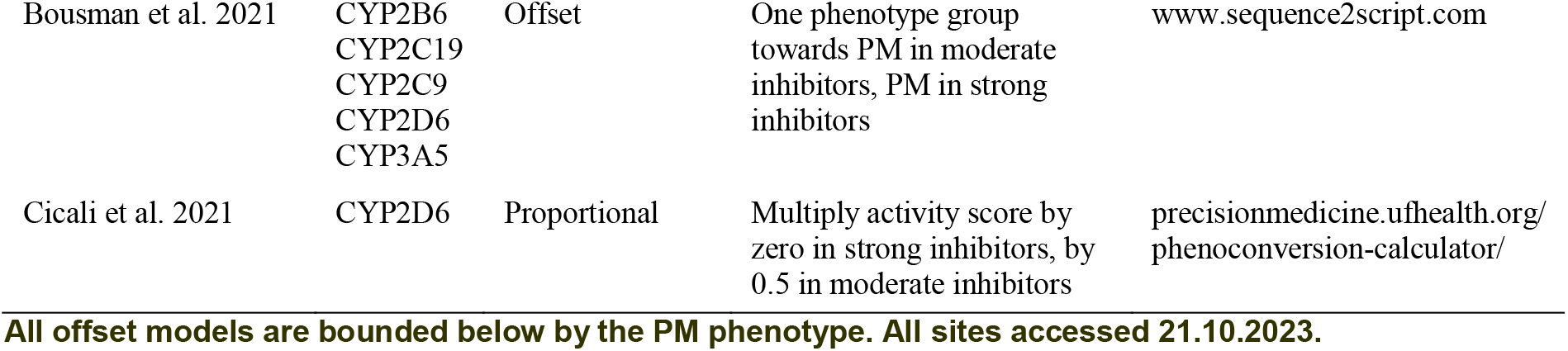
Phenotypic models of DDGI used in the literature.

### All offset models are bounded below by the PM phenotype. All sites accessed 21.10.2023

However, most of the phenotypic models in the literature are based on the suggestion of Borges et al. (2010), who evaluated two models of phenoconversion, corresponding to proportional and nonlinear phenoconversion functions, using data on the inhibition of CYP2D6 by concomitant antidepressant medication in tamoxifen-treated breast cancer patients. They concluded that the nonlinear model failed to appreciably improve the fit, advocating a proportional adjustment of activity scores by a factor of 0.5 in moderate inhibitors, and zero in strong inhibitors. This proportional model was used in most subsequent work (Lesche et al. 2020; Mostafa et al. 2021; Cicali et al. 2021), and was also adopted in CPIC guidelines (Crews et al. 2011, 2014, 2021; Hicks et al. 2016; Bousman et al. 2023).

A series of studies by the group of Michel Tod in Lyon, culminating in the systematic proposal by Fernier et al. (2018), proposed a model that stands out in that it was developed as an extension of the model by Ohno et al. (2007), thus explicitly encompassing models of both inhibition and genetic polymorphisms (Tod et al. 2013). In the framework adopted here, the phenoconversion function may be expressed as

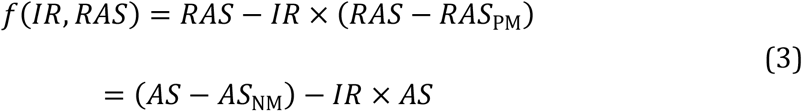

(see Appendix A3 for derivation). When there is no inhibition, *RAS* is left unaltered, and decreases proportionally to *IR* otherwise. *RAS* − *RAS*_PM_ is the activity score *AS*, which is multiplied by *IR* in the equation. From this we see that this is a proportional model.

This *linear phenoconversion* model combines the linearity of pure pharmacogenetic effects and inhibition in NMs of the previous models and contains them as particular cases. The proportional phenoconversion proposed by Borges et al. (2010) is also a particular case of this model, obtained by setting the *IR* of moderate inhibitors to 0.5 and strong inhibitors to 1. The relationship between these models is illustrated in Figure 2A.

**Figure 2.**
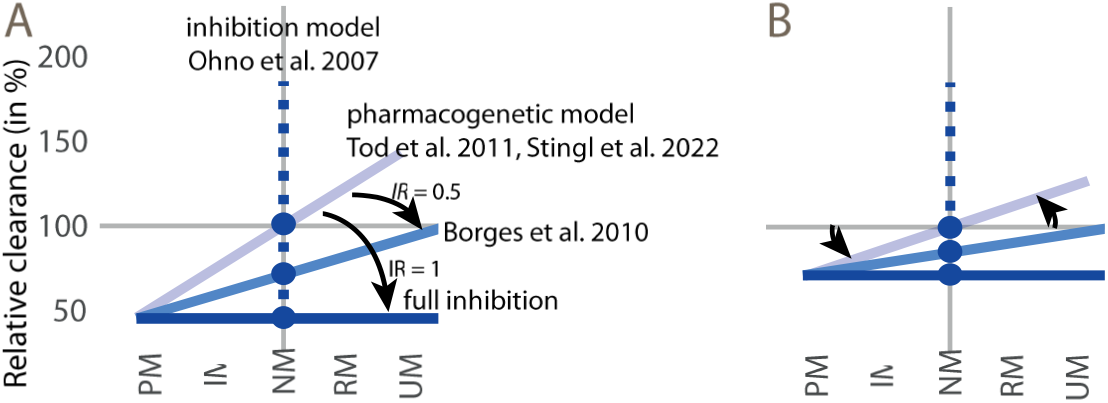
A: the linear phenoconversion model (eq. 3) includes several models as particular cases. B: effect of a reduction in the importance of the DME in the metabolic pathway. The two coefficients of the linear phenoconversion model change the slope of the linear relationship between phenotype and clearance ratio, but the changes pivot around the NM phenotype in one case and around the PM phenotype in the other.

All straight lines modelling effects of phenotype groups on clearance meet at the PM phenotype, irrespective of the extent of inhibition. The value of *IR* moves downward the slope of the straight line of the pharmacogenetic model, but such that all slopes fan out of the PM phenotype. The maximal adjustment occurs at full inhibition (*IR =* 1), in which case this line is flat at the *RAS*_PM_ level irrespective of phenotype. In NMs, where *RAS* is zero, one recovers the inhibition model of eq. 2. Figure 2B shows the effect of a change in *F*_D_.

The linear phenoconversion models is highly constrained. With only two parameters, it is possible in principle to predict DDGIs from pharmacogenetic and inhibition studies without necessarily collecting data on their joint effects (of course, data on joint effects are needed to verify the correctness of the model). Borges et al. (2010) remains the only study to have evaluated alternative models, and the results of their comparison appear to validate the linear phenoconversion model, as their proportional model is one such. However, in the presence of strong inhibitors the predictions of all models may be very similar, as the clearance is bounded below at the PM level.

In the approaches used in this literature to estimate *IR*, variations in the clinical doses of the inhibitor, which in practice constitutes co-medication, are not usually considered. Either *IR* is derived directly from clinical studies with inhibitors administered at adequate doses to exert their maximal inhibition potential (Ohno et al. 2007), or (based on the assumption that inhibition can be regarded equally to pharmacogenetic lack of enzyme activity) is estimated from pharmacogenetic studies providing the AUC or clearance difference between poor metabolizer and normal metabolizers (Tod et al. 2011).

The model assumptions we identified in this analysis are summarized in Table 2. Their implications are further discussed in the next section.

**Table 2.** Important model assumptions.

| Assumption | Model | Consequences of violation of assumptions |
| --- | --- | --- |
| Activity scores only model polymorphisms affecting DME activity | Pharmacogenetic | Polymorphisms affecting DME affinity require separate modelling |
| Polymorphisms affecting DME activity are known | Pharmacogenetic | Yet undiscovered alleles not accounted for in activity scores |
| Activity scores of genetic polymorphisms are known | Pharmacogenetic | Lack of precision in activity scores may lead to biased estimates of pharmacogenetic effects or optimistic confidence intervals of such effects |
| The fractional contribution $F_D$ of a DME in metabolism is constant across phenotypes | Pharmacogenetic | In IMs and PMs, the relation between phenotypes and clearance may be not linear in selective substrates |
| The fractional contribution $F_D$ of a DME in metabolism is constant across inhibition rates | Inhibition | The relation between inhibition and clearance may be not linear in selective substrates |
| The fractional contribution $F_D$ of a DME in metabolism is not affected by inhibition modalities | Inhibition | The fractional contribution of a DME may be altered by inhibition or induction of other DMEs |
| Phenoconversion assumption | Phenoconversion | Situation where inhibition influences $F_D$ (as in multiple inhibitors) or when pharmacogenetic variants affect inhibition properties |

### Open issues and perspectives

Pharmacogenetic models generally assume linearity in the activity scores. However, this assumption may be violated when a drug is metabolized by one DME as major metabolic pathway, because in PMs other DMEs may take over the metabolism of the drug. This causes a non-linearity in the response of low-capacity phenotypes. This situation, however, needs not invalidate the phenoconversion model. Under the phenoconversion assumption, while the relationship between activity scores and clearance is now non-linear, it may be applied to the phenoconverted activity scores as before.

Inhibition is more difficult to model than genetic polymorphism. When inhibitors affect multiple DMEs simultaneously (Isoherranen et al. 2012), it may be challenging to model the simultaneous effects. The required assumption would be that the fractional contributions of the DMEs, estimated when each was affected in isolation, apply unaltered in this situation, and that *IR* for each involved enzyme may be estimated correctly from the inhibition model.

The phenoconversion assumptions formalizes the notion that the fractional contribution *F*_D_ is not affected by inhibition and vice versa. This assumption seems reasonable in the simple situation of an inhibitor and a “victim drug”. However, the assumption may be violated in other cases. For example, inhibition may be affected by the pharmacogenetic variant itself if the inhibitor is metabolized by a polymorphic DME. Another example is reciprocal inhibition between two drugs both metabolized by the same polymorphic DMEs. The drug affinity to the enzyme and the drug dose of both substrates will determine the inhibition effect, and both drugs may partly act as perpetrator and as victim drug, creating a reinforcing loop of ihibition. These problems will require assessing the model fit empirically, and whether the phenoconversion assumption applies also in such cases. Nevertheless, the clarification of the form of the phenoconversion function in the simple situation of a strong inhibitor and a “victim drug” remains a necessary first step of any effort to build more complex interaction models.

For the purpose of individual drug dosing in polypharmacy conditions, these predictive models will require validation with pharmacokinetic real-world data. This may be a future research goal that could be pursued by creating large real-world data on drug plasma concentrations in polypharmacy conditions.

### Selection of studies

Using Google Scholar, models for drug-drug interactions were systematically investigated by looking at the literature on drug-drug-gene interactions (keyword “drug-drug-gene interaction”; PubMed gave no hits with this keyword or with “DDGI”), retrieving 345 studies. Studies were retained that included static models and mechanistic static approaches, resulting in the selection of 11 studies and 5 distinct models. Additionally, software programs and online tools for drug interactions and subsequent dose recommendations used by pharmacies, physicians and hospitals were considered and the sources of their modelling identified.

The static models thus identified were algebraically examined to identify reciprocal relationships. The analysis also considered the elementary pure pharmacogenetic or inhibition models on which the identified drug-drug-gene interaction models were based. The outcome of this analysis is detailed in the Results section, while the Appendix contains the algebraic derivations.

## Data Availability

The manuscript contains proofs of relationships between pharmacogenetic models, and the proofs are included in the manuscript

## Acknowledgments

Roberto Viviani is supported by an ERA-PERMED grant (project ArtiPro) of the FWF Austrian Science Fund (grant number I 5903). Julia Stingl is supported by an ERA-PERMED grant (project ArtiPro) of the BMBF (grant number: 01KU2212). The authors declare no conflict of interest.

## List of abbreviations

DDGI: drug-drug-gene interaction
DME: drug metabolizing enzyme
PM, IM, NM, RM, UM: poor, intermediate, normal, rapid, ultrarapid metabolizer phenotype
XM: a metabolizer phenotype variable
*AUC*_PM_, *AUC*_NM_, *AUC*_XM_, *AUC*\*: area under curve measured in PMs, NMs, in the phenotype XM, and in the presence of an inhibitor
*CL*_PM_, *CL*_NM_, *CL*_XM_, *CL*\*: clearance measured in PMs, NMs, in the phenotype XM, and in the presence of an inhibitor
*AS*: activity score of a genetic polymorphic DME, i.e. one where the value of a PM is zero
*RAS*: relative activity score of a genetic polymorphic DME, i.e. one where the value of a NM is zero
*F*_D_: a variable estimated in phenotypic models of pharmacogenetic effects, representing the importance of a DME in the metabolism of the drug *D*
*CR*: a variable in static models of inhibition representing the fraction contributed by a DME in the hepatic metabolism of a drug, being equivalent to *F*_D_ or to a rescaled version thereof
*IR*: inhibition rate, a variable representing the effect of an inhibitor on the activity of a DME ranging from zero (no inhibition) to unity (full inhibition)
*I*_app_: apparent time-averaged concentration of the inhibitor in the liver
*K*_*i*_: inhibition constant
*IX*: a variable in static models of drug-drug-gene interactions, equivalent to −*IR*.
*FA*: a variable in static models of drug-drug-gene interactions representing an activity score with value zero in PMs and unity in NMs

## Appendix

## A1. Tod’s model of effects of genotypes

Tod et al. (2011) presented a phenotypic model of drug exposure in DME genetic polymorphisms (eq. 1, p. 583). This model adapted the approach of Ohno et al. to pharmacogenetic data. The model refers to the ratio of the areas under the curve (AUC) between the genetic variant phenotype XM and the normal metabolizer NM:

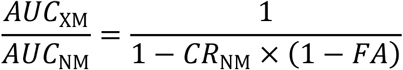

where *CR*_NM_ (called the contribution ratio) is the fraction of the apparent drug clearance due to the DME at hand, and *FA* (fractional activity) is the fraction of activity in the XM phenotype deriving from the mutated alleles, relative to the activity of the reference genotype (NM). This quantity is additive in the number of alleles, in the sense that the *FA* of an allelic composition is the average of the contribution of the individual alleles.

Rewriting in terms of clearance,

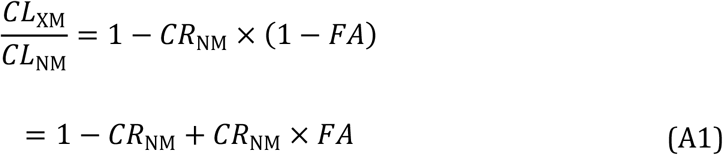

One can see that this model is linear in the fractional activity *FA*. The clearance ratio is given by the sum of a baseline term, 1 − *CR*_NM_, and a linear term, *CR*_NM_ × *FA*. Because *CR*_NM_ is a ratio (the contribution ratio of the DME), 1 − *CR*_NM_ represents the contribution ratio not due to the DME at hand. By converse, *CR*_NM_ × *FA* is the contribution of the DME, weighted by the *FA*, which is an activity score. Even if this equation has two terms, there is only one parameter given the activity score, because *CR*_NM_ appears in both terms.

### Comparison with the phenotypic model of Stingl et al. (2022)

The phenotypic model of Stingl et al. (2022) is linear and is fit by a linear regression with a constant intercept (see eq. 1 of the main text). Given the activity score, this model also has one parameter. Both models are linear but differ in that the former is expressed in terms of two ratios, *CR*_NM_ and *FA*, while in the latter *RAS* is the difference in activity of the enzyme as predicted by genotype, and *F*_D_ is estimated from the data. We now derive the relationship between these quantities.

In Tod’s model, in PMs the fractional activity *FA* is zero, as PMs have no enzymatic activity. Hence, according to eq. (A1), the clearance ratio of PMs is

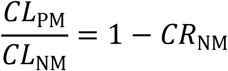

In the model of Stingl et al. (eq. 1 of the main text), let *RAS*_PM_ be the relative activity score of the PM phenotype. Then the clearance ratio of PMs is given by

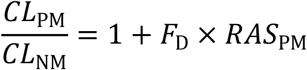

Therefore, by equating the second terms of both equations and rearranging, the contribution ratio *CR*_NM_ may be given by

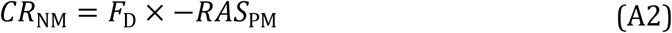

For the purpose of their use in a linear model, as the one of Stingl et al. (2022), activity scores may be rescaled without changing the fit. To gain further insight on *CR*_NM_, let the scale be that of the changes in induced by PMs, for example let a *RAS* negative unit express the contribution of two *2 alleles in CYP2C19 polymorphism. Then, when replacing *RAS*_*PM*_ with −1 in the last equation, we see that the coefficient of the linear model is on the same scale as *CR*_NM_:

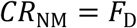

With this scaling of activity scores, *F*_D_ estimates the fractional contribution of the DME in the metabolism of the drug.

Let us now assume that the *FA*’s in Tod’s model and the *RAS*’s in Stingl’s model are known. The model by Tod et al. (eq. A1) may then be expressed in terms of *F*_D_ by replacing *CR*_NM_ according to eq. A2:

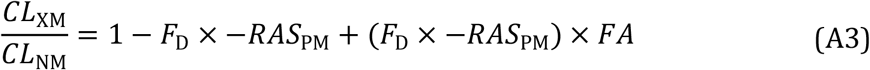

Equating with the second term of the model of Stingl et al. (eq. 1 of the main text), we obtain

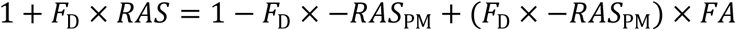

which, solving for *FA*, gives

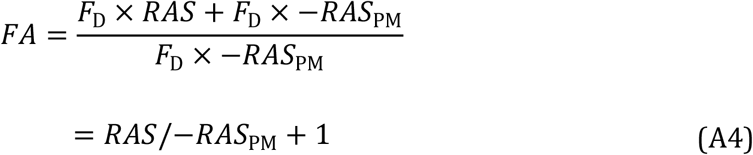

We can see here that *FA* in the model of Tod’s et al. is an activity score rescaled by minus the relative activity scores of PMs, and rebased such as to be unity in NMs.

Replacing the expression for *FA* (eq. A4) in the formula for the clearance ratio given above for the model of Tod et al. (eq. A3), one obtains

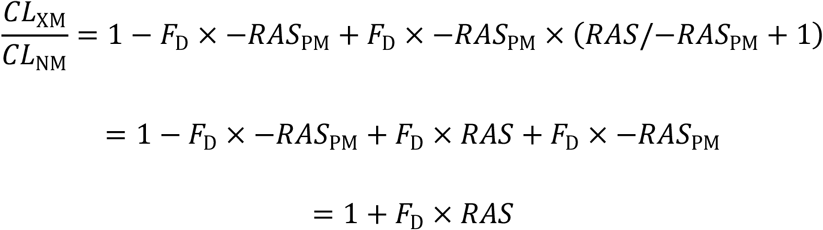

which shows that, if *FA*’s/*RAS*’s are known, the models by Stingl et al. and Tod et al. are equivalent.

The model by Tod et al. relies on data *in vivo* from normal and poor metabolizers or co-medication with inhibitors to estimate *CR*_NM_. At a second stage, an empirical Bayesian approach was used to compute confidence intervals and extend predictions of other phenotypes. Beside the different DMEs, the study of Stingl et al. (2022) differs from Tod et al. in the use of Bayesian priors to obtain prudential estimates and credibility intervals of pharmacogenetic effects.

Both models assume linear kinetic and cannot be applied if the DME contribution of other enzymes differs in PM’s and in other phenotypes.

## A2. Ohno’s model of inhibition

Denoting with an asterisk the inhibited condition, Ohno et al. (2007) proposed the following model (eq. 11, p. 46):

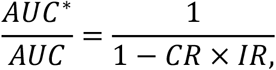

where *CR* is the fraction contributed by the DME in the hepatic metabolism of the drug, and *IR* is the inhibition rate, ranging from zero (no inhibition) to unity (full ihnibition). Expressed in terms of clearance ratios, assuming equal intestinal availability in the inhibited and unaltered condition:

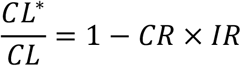

*CR* is estimated from data where drug metabolism under full inhibition or in PMs is compared to the metabolism in the reference condition (no inhibitor or NMs, as appropriate). To establish the relationship of this model to the variables of the linear model of Stingl et al. (2022), we may therefore equate clearance in the conditions of full inhibition in Ohno’s model (*IR* = 1) and clearance in PMs in the model of Stingl et al.:

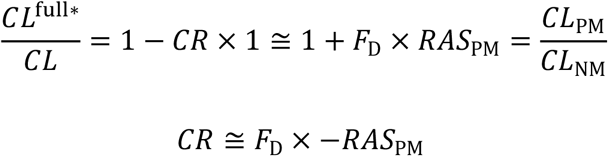

where we see that in Ohno’s model *CR* has the same relationship to *F*_D_ as *CR*_NM_ in the pharmacogenetic model by Tod et al. of the previous section (cf. eq. A2). Therefore, replacing *CR* in Ohno’s model of inhibition expressed in terms of clearance ratios, we may also write

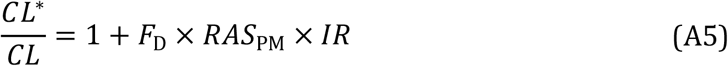

This equation says that under full inhibition (*IR* = 1), the clearance ratio is that of PMs, and when *IR =* 0 (no inhibition), the clearance ratio is unity, i.e. the clearance is not affected. Intermediate values of *IR* give clearance ratios between PMs and unity. When *F*_D_ is zero (the DME has no role in metabolizing the drug), then the clearance ratio is not altered by inhibiting the enzyme.

Note that the model is linear in *IR*. The clearance ratio decreases when inhibition is larger than zero because *RAS*_PM_ is negative. However, the clearance ratio cannot become negative because it is bounded below by the clearance ratios of PMs. Applied to an inducer, where *IR* < 0, the model can produce clearance ratios larger than unity, as it should.

To gain further insight on this model, let us encode the activity score of PMs as −1. In this case, *F*_D_ estimates the magnitude of the change in clearance ratios in PMs relative to NMs, and the above expression simplifies to

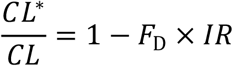

We see that here *IR* encodes the fraction of the magnitude change in PM clearance ratio that is brought about by the inhibitor.

## A3. Tod’s model of joint effects of inhibitors and genotypes

The models by Tod et al. built on the inhibition model by Ohno et al. and is therefore not surprising that notions such as *CR* have the same meaning in both. Furthermore, as we have shown in the previous sections, both models can be expressed in terms of the linear model of Stingl et al. (2022). The model for the joint effects of inhibitors and genetic polymorphisms, reported in Tod et al. (2013), should therefore be amenable to the same unified treatment. Here, we also identify its phenoconversion function.

For two DMEs, the model proposed by Tod et al. (2013) is given by (eq. 2, p. 1243)

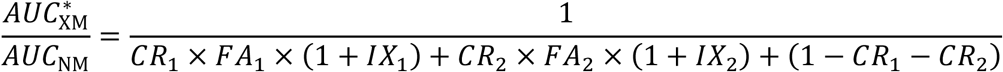

Here, the subscript refers to two DMEs, and *IX* ranges from 0 (no inhibition) to −1 (complete inhibition) and is positive in case of inducers. For inhibition, *IX* is defined analogously to Ohno’s *IR* (see Tod et al. 2013, Appendix), but is inverted in sign.

Expressing the model in terms of clearance ratios, and replacing *IX =* −*IR*, we obtain

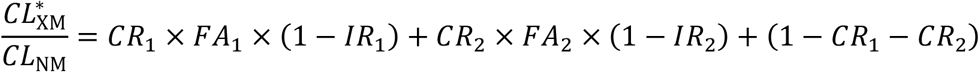

To identify the phenoconversion function implicit in this model, let us consider the case when only one enzyme is inhibited, and is the only polymorphic enzyme (i.e., *IR*_2_ *=* 0, *FA*_2_ *=* 1). Then we have

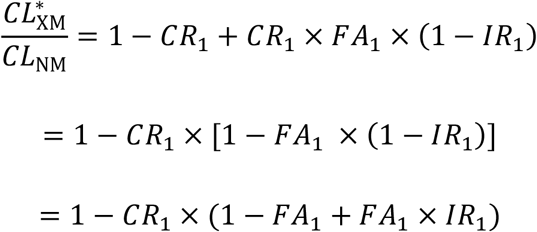

Expressed in terms of *F*_D_ and *RAS* (using eq. A2 and eq. A4), and dropping the common subscript,

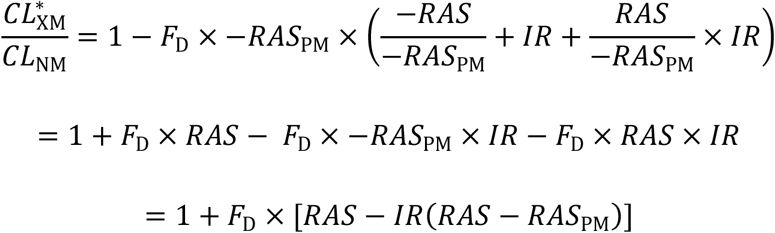

This shows that the phenoconversion function of the model is

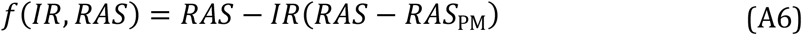

